# Immune profile of children with post-acute sequelae of SARS-CoV-2 infection (Long Covid)

**DOI:** 10.1101/2021.05.07.21256539

**Authors:** Gabriele Di Sante, Danilo Buonsenso, Cristina De Rose, Piero Valentini, Francesco Ria, Maurizio Sanguinetti, Michela Sali

## Abstract

There is increasing reporting by patients’ organization and researchers of long covid (or post-acute sequelae of SARS-CoV-2 - PASC), characterized by symptoms such as fatigue, dyspnea, chest pain, cognitive and sleeping disturbances, arthralgia and decline in quality of life. Immune system dysregulation with a hyperinflammatory state, direct viral toxicity, endothelial damage and microvascular injury have been proposed as pathologenic mechanisms. Recently, cohorts of children with PASC have been reported in Italy, Sweden and Russia. However, immunological studies of children with PASC have never been performed.

In this study, we documented significant immunologic differences between children that completely recovered from acute infection and those with PASC, providing the first objective laboratory sign of the existence of PASC in children.

## Introduction

There is increasing reporting by patients’ organization and researchers of long covid (or post-acute sequelae of SARS-CoV-2 - PASC), characterized by symptoms such as fatigue, dyspnea, chest pain, cognitive and sleeping disturbances, arthralgia and decline in quality of life^1^. Immune system dysregulation with a hyperinflammatory state, direct viral toxicity, endothelial damage and microvascular injury have been proposed as pathologenic mechanisms^1^. Recently, cohorts of children with PASC have been reported in Italy^2^, Sweden^3^ and Russia^4^. However, immunological studies of children with PASC have never been performed.

## Methods

Children with PASC after microbiologically confirmed (with PCR on nasopharyngeal swab) acute COVID-19 were identified using an internationally-developed survey (https://isaric.org/research/covid-19-clinical-research-resources/paediatric-follow-up/). PASC children were defined as those having persisting symptoms for more than five weeks and involvement of two systems or more. As a control group, we enrolled children that fully recovered from acute COVID-19. Children that fully recovered or with PASC assessed in a dedicated post-covid outpatient service were invited to perform an immunological assessment which included:

- T regulatory and B cells subsets, from freshly collected blood samples, labelled with DuraClone^®^ lyophilized antibody panels and analysed with Kaluza^®^ 2.1 (Beckman Coulter, Pasadena, CA, USA).
- expression levels of cytokines IL6, IL1β and TNFα, using the ELLA system (ProteinSimple, San Jose, California, USA) according to the manufacturer’s protocol.

The study was approved by the ethic committee of our Institution (ID 3078). Written informed consent was obtained from all participants or legal guardians.

## Results

We enrolled 12 children with PASC (long covid) and 17 that completely recovered from the acute infection (Table 1).

**Table 1.** Characteristics of children that fully recovered from acute infection and those with Post-Acute sequelae of SARS-CoV-2 (PASC) (also known as long covid). Pre-existing conditions: Recovered: Obesity (1), allergic asthma (1); recurrent respiratory tract infections (1); Intraventricular Defect operated at one year of age (1). PASC: allergic asthma (1); past diagnosis of Schönlein-Henoch Purpura (1); Duchenne with no oxygen support, which suffered, after mild acute COVID-19, of headache, palpitations and an unexplained 20% reduction of ejection fraction. *For acute pediatric COVID-19 the following classification was used (adapted by first Italian and Chinese studies revised at N Engl J Med. 2020 Jul 9;383(2):187-190. doi: 10.1056/NEJMc2007617): -Asymptomatic infection: without any clinical symptoms and signs, nor abnormal radiologic findings -Mild: symptoms of acute upper respiratory tract infection, including fever, fatigue, myalgia, cough, sore throat, runny nose, and sneezing. -Moderate: with pneumonia, frequent fever, and cough (mostly dry cough, followed by productive cough) -Severe: early respiratory symptoms, such as fever and cough, may be accompanied by gastrointestinal symptoms, such as diarrhea. Oxygen saturation is <92% with other hypoxia manifestations.

|  | **Fully**<br>**Recovered**<br>**N 17 (%)** | **Long covid /**<br>**PASC**<br>**N 12 (%)** |
| --- | --- | --- |
| **Female, n° (%)** | 6 (35.5) | 4 (33.3) |
| **Age, mean ± SD** | 7.7 ± 5.5 | 10.3 ± 4.5 |
| **Acute COVID-19 severity\*, n° (%)** |  |  |
| *Asymptomatic* | 2 (11.8) | 1 (8.3) |
| *Mild* | 10 (58.8) | 11 (91.7) |
| *Moderate* | 3 (17.6) | / |
| *Severe* | 2 (11.8) | / |
| **Hospitalized during acute COVID-19, n° (%)** | 5 (29.4) | 0 |
| **Days after acute SARS-CoV-2 infection, mean ± SD** | 101 ± 40.8 | 98.5 ± 41.5 |
| **Persisting Symptoms, n° (%)** | / | 12 (100) |
| *Fatigue* | / | 5 (41.6) |
| *Chronic Headache* | / | 4 (33.3) |
| *Gastrointestinal symptoms* | / | 4 (33.3) |
| *Post-exertional malaise* | / | 3 (25.0) |
| *Muscle or Joint pain* | / | 3 (25.0) |
| *Chest pain* | / | 3 (25.0) |
| *Low grade fever* | / | 2 (16.6) |
| *Chronic cough* | / | 2 (16.6) |
| *Tachicardia* | / | 1 (8.3) |
| *Sleeping disorders* | / | 1 (8.3) |
| **Pre-existing conditions, n° (%)** | 3 (17.6) | 3 (25.0) |

Distinct immunologic features were detectable comparing PASC patients and children who had recovered. The group of PASC showed significantly higher levels of plasmablasts, IgD^-^CD27^+^ memory and switched IgM^-^IgD^-^ B cells. On the contrary, healed children had significantly higher naïve and unswitched IgM^+^IgD^+^ and IgM^+^CD27^-^CD38^dim^ B cell subsets (figure 1a). The T-regulatory compartment showed no significant differences (Figure 1b). Moreover, IL6 and IL1β serum levels were elevated in PASC patients and consistently higher than children who had recovered after infection (Figure 1c).

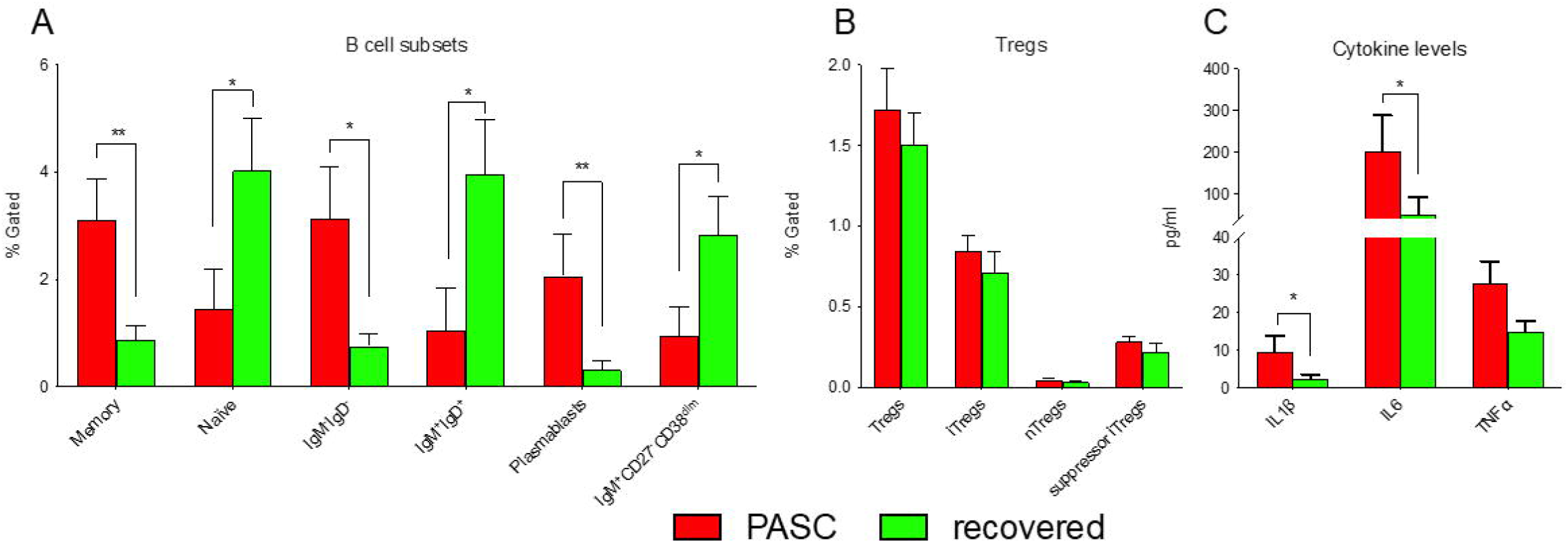
Cellular and cytokine immune profile of PASC and fully recovered children. Unpaired multiple comparison between PASC (red bars) and fully recovered children (green bars) were performed with GraphPad 9.0 software (Prism) and Mann-Whitney test, corrected using Holm-Šidàk method. Details pertaining to significance are noted. A) The staining with CD45-KrO, CD19-ECD, CD27-PC7, IgD-FITC, CD38-APC750, CD21-PE and IgM-PB revealed in PACS than in recovered children higher amounts of plasmablasts, IgD^-^CD27^+^ memory and switched IgM^-^IgD^-^ B cells (2.06%±2.74 vs 0.31%±0.74, p=0.008; 3.11%±2.66 vs 0.89%±1.19, p=0.005; 3.12%±3.41 vs 0.76%±0.96, p=0.02, respectively) and lower percentages of naïve and unswitched IgM^+^IgD^+^ and IgM^+^CD27^-^CD38^dim^ B lymphocytes (1.45%±2.6 vs 4.04%±3.97, p=0.02; 1.06%±2.71 vs 3.97%±4.16, p=0.02; 0.95%±1.84 vs 2.82%±3.03, p=0.02, respectively). B) T regulatory lymphocytes, labelled with CD45-KrO, CD4-PC7, CD3-APC750, CD25-PE, FoxP3-A647, Helios-PB, CD39-PC5.5, CD45RA-FITC, did not show significant differences between the two groups. C) The comparison of pro-inflammatory cytokines shows significant higher levels of IL1β and IL6 in the sera of PASC than of fully recovered children (9.58±14.4 vs 2.36±4.72, p=0.05; 202.73±297.4 vs 50.02±173.45, p=0.03) and a trend for TNFα (27.7±20.67 vs 15.02±10.25, p=0,06).

## Discussion

While PASC is widely recognized in adults^1^, its existence in children is more controversial, since it has been speculated that this group may rather have psychological sequelae of social restrictions. In this study, we documented significant immunologic differences between children that completely recovered from acute infection and those with PASC, providing the first objective laboratory sign of the existence of PASC in children. Healed children restored B-cell homeostasis, by reconstituting the naïve/unswitched B-cell compartment. In PASC children, the consistent amounts of plasmablasts, switched and memory B lymphocytes revealed the involvement of those subsets in the immunopathogenesis of chronic symptoms. Moreover, the persisting high levels of IL6 and IL1β suggest that innate immune response played a pivotal role in children with PASC. Considering their role as mediators of inflammatory responses and auto-immune processes, these findings could explain systemic persisting symptoms such as fatigue and post-exertional malaise, headache, muscle and joint pain, and tachycardia.

The possibility that a low level, persistent infection after acute infection may persist in some children, contributing to the development of post-acute inflammatory conditions such as Multisystem Inflammatory Syndrome (MIS-C) or PASC has recently been suggested. The presence of persisting viral particles were recently detected by immune-histochemistry and electron microscopy in dermal vascular endothelium weeks after acute infection^5^. Moreover, extensive burr cells (echinocytes) have been described in the blood smears of patients with MIS-C (which typically develops weeks after acute infection), and have been linked to inflammatory conditions^6^. Our findings provide further evidence that a chronic inflammatory process and immune dysfunction may explain why some children develop PASC, highlighting the importance of further studies aimed to better characterize this new condition. A better understating of PASC will also provide indirect benefits for other post-infectious conditions.

Limitations of our study are the low number of children included and the lack of ex-vivo immunologic studies performed after in-vitro stimulation with SARS-CoV-2, which we plan as a future study.

## Data Availability

dataset is available upon request to the corresponding author

## Notes

**Conflict of interests:** nothing to declare

### Competing Interest Statement

The authors have declared no competing interest.

### Funding Statement

nothing to declare

### Author Declarations

Comitato Etico Fondazione Universitario Policlinico A. Gemelli

## References

1. Nalbandian A, Sehgal K, Gupta A, et al. Post-acute COVID-19 syndrome. Nat Med. 2021 Apr;27(4):601–615. doi: 10.1038/s41591-021-01283-z

2. Buonsenso D, Munblit D, De Rose C et al. Preliminary evidence on long COVID in children. Acta Paediatr. 2021 Apr 9. doi: 10.1111/apa.15870

3. Ludvigsson JF. Case report and systematic review suggest that children may experience similar long-term effects to adults after clinical COVID-19. Acta Paediatr. 2021 Mar;110(3):914–921. doi: 10.1111/apa.15673.

4. Osmanov IM, Spiridonova E, Bobkova P, et al. Risk factors for long covid in previously hospitalised children using the ISARIC Global follow-up protocol: A prospective cohort study. medRxiv 2021.04.26.21256110. doi:10.1101/2021.04.26.21256110

5. Colmenero I, Santonja C, Alonso-Riaño M, et al. SARS-CoV-2 endothelial infection causes COVID-19 chilblains: histopathological, immunohistochemical and ultrastructural study of seven paediatric cases. Br J Dermatol. 2020 Oct;183(4):729–737. doi: 10.1111/bjd.19327.

6. Diorio C, Henrickson SE, Vella LA, et al. Multisystem inflammatory syndrome in children and COVID-19 are distinct presentations of SARS-CoV-2. J Clin Invest. 2020 Nov 2;130(11):5967–5975. doi: 10.1172/JCI140970

